# Long-read metagenomics reveals a high burden of antimicrobial resistance, mobile genetic elements and bacterial diversity in hospital and community wastewater from Conakry, Guinea

**DOI:** 10.64898/2026.08.14.26360450

**Authors:** Thibaut Armel Chérif Gnimadi, Alpha Kabiné Keita, Yaovi Mahuton Gildas Hounmanou, Kekeli Elodie Awounon, Jean-François Zagury, Abdoulaye Toure, Mano Joseph Mathew, Alpha Kabinet Keita

**Affiliations:** Centre de Recherche et de Formation en Infectiologie de Guinée (CERFIG), Université Gamal Abdel Nasser de Conakry, Conakry 6629, Guinea; Laboratoire Génomique, Bioinformatique et Chimie Moléculaire, EA7528, Conservatoire National des Arts et Métiers, HESAM Université, 2 Rue Conté, 75003 Paris, France; Department of Veterinary and Animal Sciences, Faculty of Health and Medical Sciences, University of Copenhagen, Frederiksberg, Denmark; EFREI Research Lab, Panthéon Assas University, 30-32 Avenue de la République, 94800 Villejuif, France; Institut de Recherche pour le Développement (IRD), INSERM, TransVIHMI, University of Montpellier, 34394 Montpellier, France

**Keywords:** AMR, Resistance Genes, Microbiome, Wastewater, Long-read, Nanopore, Genome-resolved metagenomes, Guinea

## Abstract

Wastewater systems are increasingly recognized as important environmental reservoirs of antimicrobial resistance (AMR), acting as interfaces where resistant bacteria, antimicrobial resistance genes (ARGs), and mobile genetic elements (MGEs) converge and potentially disseminate. Wastewater samples were collected from hospital and community sites, including municipal medical centers, household wastewater outlets, and open drainage systems. Genomic DNA was extracted using the ZymoBIOMICS™ DNA/RNA Miniprep Kit and sequenced on the Oxford Nanopore Technologies MinION MK1D platform using the Native Barcoding Kit (SQK-NBD114.24, V14). Sequencing data were processed through a custom Snakemake workflow integrating quality control, taxonomic profiling, resistome characterization, mobilome analysis, and genome-resolved metagenomics. A total of 489 unique ARGs conferring resistance to 29 antibiotic classes were identified through metagenomic analysis. The resistome was dominated by genes conferring resistance to β-lactams (including cephalosporins and carbapenems), aminoglycosides, tetracyclines, macrolides, and fluoroquinolones. Clinically important resistance determinants, including *bla*OXA, *bla*TEM, *bla*GES, *bla*CARB, *cfxA, tet, qnr, sul, dfrA, erm, msrE*, and aminoglycoside-modifying enzyme genes such as aac(3) and ant(3’’) were detected across both hospital and community wastewater samples. Resistance mechanisms were predominantly driven by antibiotic inactivation, followed by efflux and target protection. Several priority bacterial pathogens were detected, including *Escherichia coli, Klebsiella pneumoniae, Enterobacter cloacae, Pseudomonas aeruginosa,* and *Acinetobacter baumannii*. Integration/excision elements were the predominant category of MGEs, followed by transfer-associated elements and replication/recombination/repair functions. Plasmid analysis further identified diverse incompatibility groups, predominantly IncP6, IncC, IncF, and IncR replicons, highlighting the widespread occurrence of plasmid-associated genetic mobility in both settings. Genome-resolved analysis reconstructed 103 dereplicated MAGs, of which 80 carried ARGs and 26 contained putative mobile resistance regions defined by ARG–MGE co-localizations within 10 kb. These findings reveal a substantial burden of clinically relevant ARGs, mobile genetic elements, and potential bacterial pathogens in hospital and community wastewater in Conakry. This study provides the first metagenomic baseline for environmental AMR surveillance in Guinea and highlights the urgent need for integrated One Health strategies to mitigate the environmental dissemination of antimicrobial resistance.

## 1. Introduction

Antimicrobial resistance (AMR) is recognized as one of the greatest global public health threats of the twenty-first century. In 2019 alone, bacterial AMR was directly responsible for an estimated 1.27 million deaths and associated with nearly 4.95 million deaths worldwide, with the highest burden occurring in low– and middle-income countries, particularly in sub-Saharan Africa [1]. In addition, the continued emergence and spread of multidrug-resistant pathogens threaten the effectiveness of antimicrobial therapy and compromise the management of common infectious diseases [2]. Although AMR has traditionally been considered primarily from a human-health perspective, increasing evidence indicates that environmental compartments play a critical role in the maintenance and dissemination of resistance determinants [3].

Within the One Health framework, wastewater systems represent a major interface linking human populations, animals, and the environment, where resistant microorganisms, antimicrobial residues, heavy metals, biocides, and other pollutants converge [3,4]. Therefore, the high microbial density and taxonomic diversity of wastewater create the ideal conditions for horizontal gene transfer via conjugation, transformation, and transduction, facilitated by mobile genetic elements including plasmids, integrons, insertion sequences, and transposons [5].

Hospital and community wastewaters are particularly relevant in this context. Hospital wastewater (HWW) receive large quantities of antibiotics, disinfectants, and resistant microorganisms originating from patients undergoing antimicrobial treatment [6,7], whereas municipal wastewater reflects community-level antibiotic consumption and microorganisms shed by the human population [8]. Together, these systems constitute a continuum through which resistant bacteria and ARGs can enter natural aquatic ecosystems and potentially re-enter human and animal populations [9].

Metagenomic investigations have revealed that wastewater environments harbor a highly diverse resistome, often containing hundreds of unique ARGs belonging to nearly all clinically relevant antibiotic classes [7,10,11]. The recurrent detection of clinically relevant ARGs that confer resistance to critically important antibiotics, including carbapenemases (*blaNDM, blaKPC, blaOXA, blaGES*, and *blaVIM*) and mobile colistin resistance genes (*mcr*), is a cause for particular concern [7,12]. These resistance determinants are frequently associated with opportunistic and pathogenic bacteria of major public health importance, including members of the ESKAPE group such as *Klebsiella pneumoniae*, *Pseudomonas aeruginosa*, and *Acinetobacter baumannii* [4,13–15].

The dissemination potential of wastewater resistomes largely depends on their mobility, facilitated by MGEs and determines their capacity to spread across bacterial populations. Plasmids, transposons, integrons, bacteriophages, and integrative conjugative elements (ICEs) mediate horizontal gene transfer (HGT), enabling the rapid dissemination of resistance determinants among phylogenetically distant microorganisms [3,16].

In low-resource settings, particularly in West African countries, where sanitation infrastructure remains limited and environmental surveillance systems are often underdeveloped, studies have reported the presence of clinically relevant ARGs in untreated or poorly managed HWW, highlighting the environmental dissemination of resistance determinants in urban settings [12,17,18].

Despite increasing recognition of wastewater as a critical environmental reservoir of antimicrobial resistance, metagenomic surveillance remains extremely limited across most West African countries. In Guinea, information regarding wastewater microbial communities, ARG diversity, and MGEs remains largely unavailable. Moreover, few studies from the West Africa region have applied long-read metagenomic sequencing, which enables a more comprehensive characterization of resistance determinants and their genetic context than conventional approaches.

To address these knowledge gaps, we applied MinION Mk1D Oxford Nanopore long-read metagenomic sequencing to characterize wastewater collected from hospital and municipal sources in Conakry, Guinea. Specifically, this study aimed to (i) characterize the bacterial community structure, (ii) investigate the diversity and abundance of antimicrobial resistance genes, (iii) describe the distribution of mobile genetic elements associated with resistance dissemination, and (iv) compare hospital and community wastewater as potential environmental reservoirs of AMR. This study provides the first long-read metagenomic characterization of hospital and municipal wastewater in Guinea and contributes baseline evidence for integrating environmental surveillance into national One Health AMR monitoring programs.

## 2. Materials and Methods

### 2.1 Study Design, Sample Collection and Processing

Hospital wastewater samples were collected from sewage discharge systems and septic tanks of four secondary-level healthcare facilities, designated H1–H4. These municipal medical centers are geographically distributed across the city of Conakry and contribute to provide healthcare services to a general population of 3.4 million. Hospital wastewater samples were collected during two sampling campaigns: the rainy season in July 2025 (H_R) and the dry-to-transition season in May 2026 (H_D). One sample (H1_D) were excluded during the second sampling campaign due to the inaccessibility of wastewater sample site.

Community wastewater samples were collected from eight distinct locations (C1–C8) distributed across several urban districts of Conakry. Sampling sites included household wastewater discharge points and urban municipal open drainage channels wastewater, and they were selected on the basis of their accessibility and the high risk of contact with people and animals living in the surrounding area.

All wastewater samples were collected as grab samples using a sterile long-handled sampling device and transferred into sterile wide-mouth polypropylene containers (250 mL). Sampling was performed during mid-morning hours (08:00–11:00 a.m.) to minimize temporal variability associated with wastewater flow patterns. No regular wastewater treatment plants were available, and we didn’t consider whether wastewater was treated or not. All sample were collected at same depth of approximately 1 meter. All community sites are located some distance from hospital sites, thereby minimising the risk of hospital waste contaminating community wastewater management systems.

Immediately after collection, samples were placed in insulated coolers containing ice packs and transported to the laboratory within 1 hour while maintaining a temperature of approximately 4°C. For each sample, associated metadata were recorded, including sample identification code, source category (hospital or community), collection date and time, geographic coordinates (latitude and longitude), and relevant information regarding the sampling site.

Upon arrival at the laboratory, samples were homogenized by vigorous mixing by vertexing. An aliquot of 50 mL was transferred into sterile centrifuge tubes and centrifuged at 5,000 × g for 10 min. Following centrifugation, the supernatant was carefully discarded, and the resulting pellet was retained for subsequent genomic DNA extraction.

### 2.2 Genomic DNA Extraction and Shotgun Metagenomic Sequencing

Genomic DNA was extracted from the pellets obtained after centrifugation of wastewater samples using the ZymoBIOMICS™ DNA/RNA Miniprep Kit (Zymo Research, Irvine, CA, USA), following the manufacturer’s protocol. DNA concentration was measured using a Qubit® fluorometer (Thermo Fisher Scientific, Waltham, MA, USA) with the Qubit™ 1X dsDNA High Sensitivity (HS) Assay Kit. Quality and integrity of the DNA were assessed by electrophoresis on a 1% agarose gel.

Shotgun metagenomic libraries were prepared using a PCR-free workflow with the Ligation Sequencing Kit coupled with the Native Barcoding Expansion 24 V14 (SQK-NBD114.24; Oxford Nanopore Technologies, Oxford, UK). For each sample, approximately 400 ng of genomic DNA was used as input for library preparation. Following library construction, the pooled library was quantified using the Qubit™ dsDNA High Sensitivity Assay Kit. The final library was loaded onto FLO-MIN114 R10.4.1 flow cells and sequenced on a MinION Mk1D device (Oxford Nanopore Technologies). All runs were processed for 48h to 72h.

### 2.3 Bioinformatics and Statistical Analyses

Raw nanopore sequencing data were basecalled in high-accuracy mode (HAC) and demultiplexed using Dorado (v0.9.5). Adapter trimming, quality filtering, and read preprocessing were performed using Fastplong [19]. To remove potential host contamination, filtered reads were aligned against the human reference genome (GRCh38_no_alt) using Minimap2, and human-derived reads were removed using Samtools [20,21].

Taxonomic profiling was performed on quality-filtered reads using Kraken2 (v2.1.7) and Bracken (v3.0.1) against the standard Kraken2 database [22,23].

Taxonomic richness saturation was evaluated using rarefaction curve analysis based on multinomial subsampling with 30 iterations per sequencing depth. To account for differences in sequencing output among samples, taxonomic abundances were expressed as gigabase-weighted relative abundances (%). Bracken read counts were converted to nucleotide abundances using the average read length estimated from NanoPlot statistics and subsequently normalized by the total sequencing output of each sample.

ARGs were identified directly from metagenomic reads using the Resistance Gene Identifier (RGI bwt) workflow and the Comprehensive Antibiotic Resistance Database (CARD, v3.2.7) [24]. Detected ARGs were subsequently classified according to their associated antimicrobial classes and resistance mechanisms.

To characterize the assembly-based MGEs and reconstruct MAGs, metagenomic assemblies were generated independently for each sample using Flye v2.9.6 with the –meta and –nano-hq parameters optimized for long-read metagenomic datasets [25]. Assembled contigs were polished using medaka.

For genome-resolved metagenomic analysis, quality-filtered reads from each sample were mapped back to their corresponding assemblies using Minimap2 (v 2.28), and alignment files were processed with Samtools (v1.21) to estimate contig coverage. Metagenome binning was performed independently using MetaBAT2 (v 2.17) and SemiBin2 (v 2.3.0), and the resulting bins were integrated with DAS Tool (v 1.1.7) to generate a consensus set of MAGs. MAG quality was assessed using CheckM2 (v1.1.0), and genomes with completeness ≥50% and contamination ≤10% were retained for downstream analyses. Redundant MAGs were dereplicated using dRep (v3.5.0) based on average nucleotide identity (ANI). Taxonomic classification of MAGs was performed using GTDB-Tk against the Genome Taxonomy Database (GTDB, release 232) [26,27]. ARGs were identified within MAGs using the Resistance Gene Identifier (RGI main) against CARD, v3.2.7.

Assembled contigs and MAGs were aligned against the mobileOG-db database (data version: Beatrix) using DIAMOND (v 0.8.36) [28]. Identified MGEs were categorized into five major functional groups: Replication/Recombination/Repair (RRR), Integration/Excision (IE), Stability/Transfer/Defense (STD), Inter-Organism Transfer (T), and Phage-related biological processes (P). Plasmid replicons were identified among assembled contigs using PlasmidFinder (v 2.1.6).

Putative mobile resistance regions were defined as contigs containing at least one ARG and one MGE located within a maximum distance of 10 kb. Co-localization analyses were performed by integrating ARG annotations, mobileOG-db hits, and MAG assignments.

For ARG and MGE abundance comparisons, normalized copy numbers were calculated using the following formula:

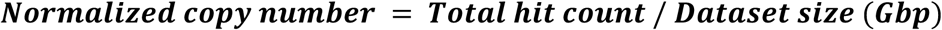

where dataset size (Gbp) was calculated as the total nucleotide yield of each sequencing library divided by 10⁹. This normalization approach enabled quantitative comparisons of ARG and MGE abundances across samples with varying sequencing depths.

Microbial alpha and beta diversity based on genus-level taxonomic data were assessed using the R packages phyloseq (v1.44) and vegan (v2.6-4). Diversity metrics included the Shannon (H′) and Simpson (1−D) indices, calculated for both bacterial genus-level composition and ARG abundance; differences between hospital and community wastewater were assessed using the Mann–Whitney U test. Differential abundance of individual taxa was assessed on CLR-transformed data restricted with FDR correction (Benjamini-Hochberg, q < 0.05). Beta diversity was assessed using Aitchison distance (CLR-transformed genus abundance) for the bacterial community and Bray-Curtis dissimilarity for the resistome. Group differences were tested using PERMANOVA (999 permutations). Because hospital sites were sampled during two seasonal campaigns while community sites were sampled once, all diversity comparisons were repeated separately for each campaign (H_R, n=4; H_D, n=3) as a sensitivity analysis alongside the pooled dataset.

All analyses were implemented within a reproducible Snakemake workflow integrating quality control, taxonomic profiling, resistome characterization, mobilome analysis and plasmid detection. The Detailed Snakemake workflows are found in https://github.com/armelgnimadi/Resistome

## 3. Results

A total of 15 wastewater samples were collected from 12 sampling sites distributed across Conakry, Guinea, including eight community wastewater sites (C1–C8) and four hospital wastewater sites (H1–H4). **(Figure 1)**

**Figure 1.**
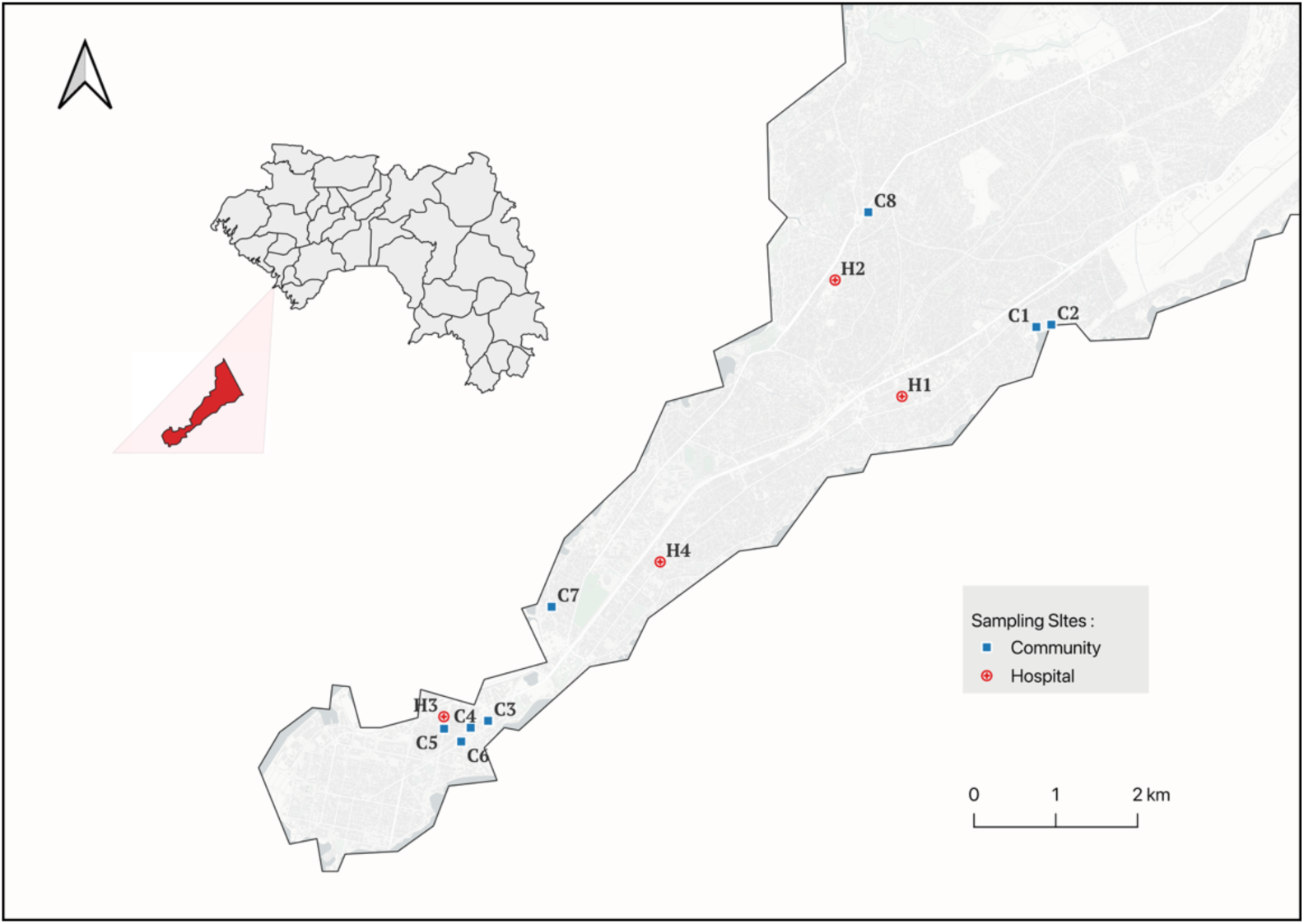
Geographic distribution of the wastewater sampling sites included in this study.

The study generated a total of 25.84 gigabases (Gbp) of sequencing data across the 15 samples, comprising 16.76 million reads, with a mean quality score of 15.6 and an average read length of 2.01 kilobases per sample **(Supplementary table S1)**

### 3.1 Microbial diversity in hospital and community wastewater

At the phylum level, bacterial communities were dominated by Pseudomonadota, Bacillota, and Bacteroidota, which together accounted for 83.3% of the overall relative abundance. Pseudomonadota was the most abundant phylum overall (42.62%), showing a higher mean abundance in community than hospital wastewater (54.13% vs. 29.47%); however, this difference did not reach significance (Mann-Whitney U q=0.279), nor did the apparent difference in Bacillota abundance (17.45% vs. 28.06%; q=0.265). In contrast, Bacteroidota was significantly enriched in hospital wastewater (29.23% vs. 8.63% in community; q=0.033), and Thermodesulfobacteriota was significantly enriched in community wastewater, with an approximately ten-fold difference (6.04% vs. 0.61%; q=0.003). Methanobacteriota and Actinomycetota showed no significant inter-source variation (q=0.343 and q=0.297, respectively). **Figure 2A**

**Figure 2.**
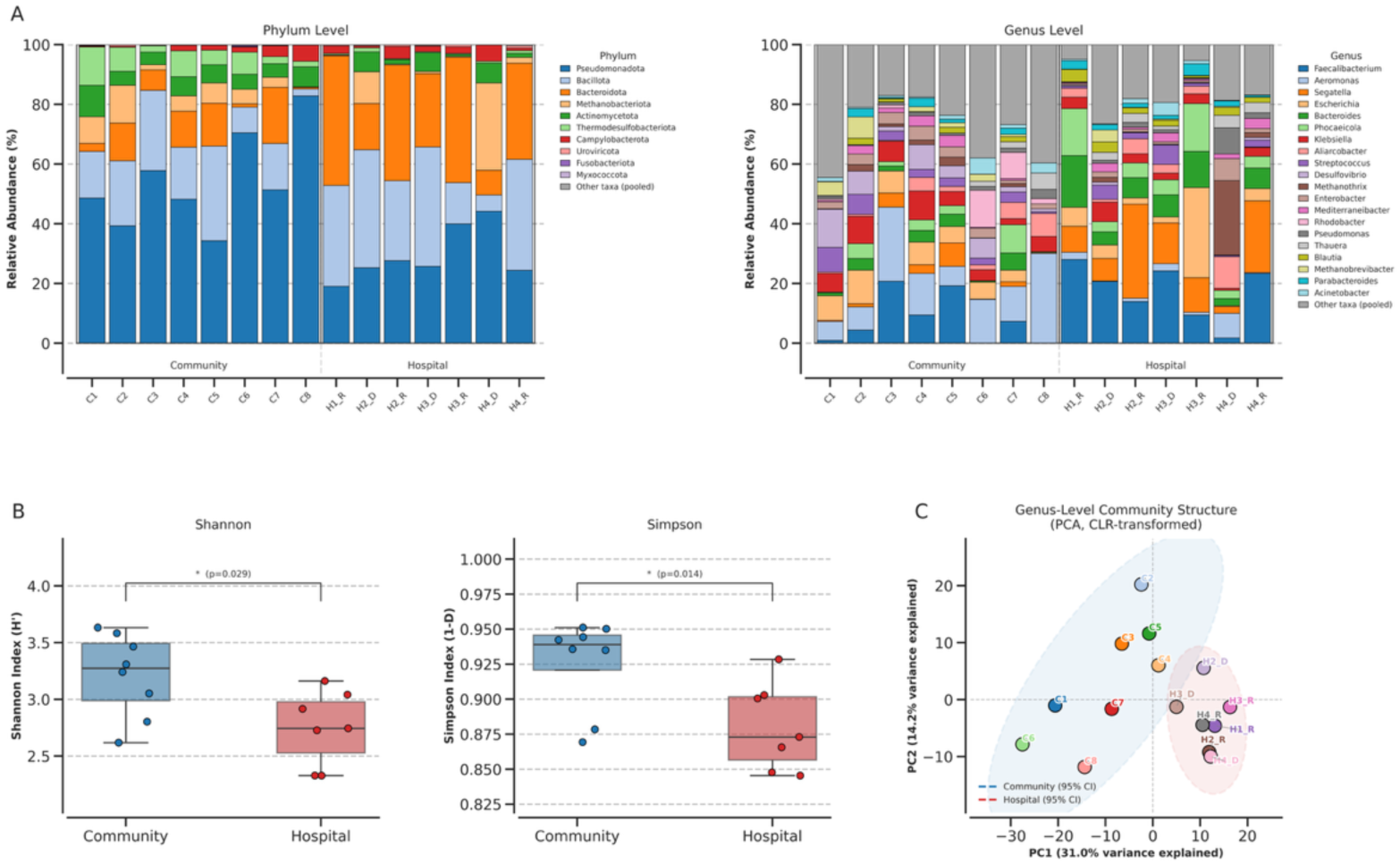
Bacterial community composition and diversity between hospital and urban community wastewater. 2A: Relative abundance of the top bacterial phyla and Genus identified across hospital and community wastewater samples 2B: Alpha diversity analysis (Shannon and Simpson indices) of bacterial composition comparing urban community and hospital sources. 2C: Principal coordinate analysis (PCoA) of genus-level bacterial composition. Community structure differed significantly between sample types (PERMANOVA, Pseudo-F = 4.12, R² = 0.240, p = 0.001; 999 permutations), while multivariate dispersions were homogeneous (PERMDISP, p = 0.307).

At the genus level, the microbiome was characterized by a mixture of fecal-associated, environmental, and clinically relevant taxa. The most abundant genera were Faecalibacterium (12.26%), Aeromonas (8.75%), Segatella (7.85%), Escherichia (6.62%), Bacteroides (5.22%), Phocaeicola (4.96%), and Klebsiella (4.66%). Differential abundance testing confirmed significantly higher abundances in hospital wastewater for Faecalibacterium (17.37% vs. 7.78%; q=0.035), Segatella (14.15% vs. 2.32%; q=0.022), Bacteroides (8.16% vs. 2.64%; q=0.035), and Phocaeicola (7.40% vs. 2.82%; q=0.045). Aeromonas showed a higher mean abundance in community wastewater (14.46% vs. 2.23%) but did not reach significance after correction (q=0.117). **Figure 2A**

Read-based species-level profiling identified several clinically relevant bacterial species in both community and hospital wastewater. *Escherichia coli, Klebsiella pneumoniae, Enterobacter cloacae, Pseudomonas aeruginosa, and Acinetobacter baumannii* were detected in all samples. *E. coli* was among the most abundant clinically relevant species (6.79%), followed by *K. pneumoniae (*4.57%), *E. cloacae* (2.05%), *P. aeruginosa* (1.29%), and *A. baumannii* (0.29%). Other pathogens, including *Clostridioides difficile, Clostridium perfringens, Enterococcus faecium, Salmonella enterica, Vibrio cholerae, and Staphylococcus aureus*, were also detected at lower relative abundances. **(Supplementary Data S2)**

Alpha diversity analysis revealed significantly higher bacterial diversity in community wastewater compared with hospital wastewater. The Shannon diversity index was significantly greater in community samples (p = 0.029), indicating higher taxonomic richness and evenness. Similarly, Simpson diversity values were significantly higher in community wastewater (p = 0.014), suggesting a more balanced community structure **(Figure 2B).**

Our sensitivity analysis comparing community wastewater separately against each hospital campaign show that the difference with the rainy-season campaign (H_R, n=4), in bacterial diversity remained significant (Shannon: p=0.016; Simpson: p=0.008). Against the dry-season campaign (H_D, n=3), the difference was no longer statistically significant (Shannon: p=0.376; Simpson: p=0.279).

Consistent with the pooled alpha diversity findings, principal coordinate analysis based on Aitchison distance show a separation between community and hospital wastewater microbiomes. PERMANOVA confirmed that wastewater source significantly influenced bacterial community composition (Pseudo-F = 4.12, R² = 0.240, p = 0.001), explaining approximately 24% of the observed variance. The assumption of homogeneous multivariate dispersions was met (PERMDISP, p = 0.307), supporting the validity of the PERMANOVA results **(Figure 2C)**. The separation between community and hospital microbiomes remained statistically significant against both the rainy-season (H_R: Pseudo-F=3.85, R²=0.278, p=0.004) and dry-season campaigns (H_D: Pseudo-F=2.21, R²=0.197, p=0.018).

### 3.2 Diversity of antimicrobial resistance genes in hospital and community wastewater

A total of 489 unique antimicrobial resistance genes (ARGs) were identified across all wastewater samples. Among these, 368 ARGs were detected in community wastewater and 359 ARGs in hospital wastewater, with 238 ARGs shared between both environments. Normalized ARG abundance was comparable between community and hospital wastewater (838.6 vs. 837.4 copies/Gb per sample; p = 0.955).

The heatmap of the 50 most abundant ARGs revealed a broad distribution of resistance determinants across all samples. Several ARG families were consistently detected in both community and hospital wastewater, forming a common wastewater resistome dominated by genes associated with resistance to β-lactams, aminoglycosides, tetracyclines, nitroimidazoles, and multidrug resistance. Among the most abundant ARGs were members of the *bla*CfxA, *bla*TEM, *blaOXA*, *aad, aac, aph, tet*, and *nim* gene families (**Figure 3A)**.

**Figure 3:**
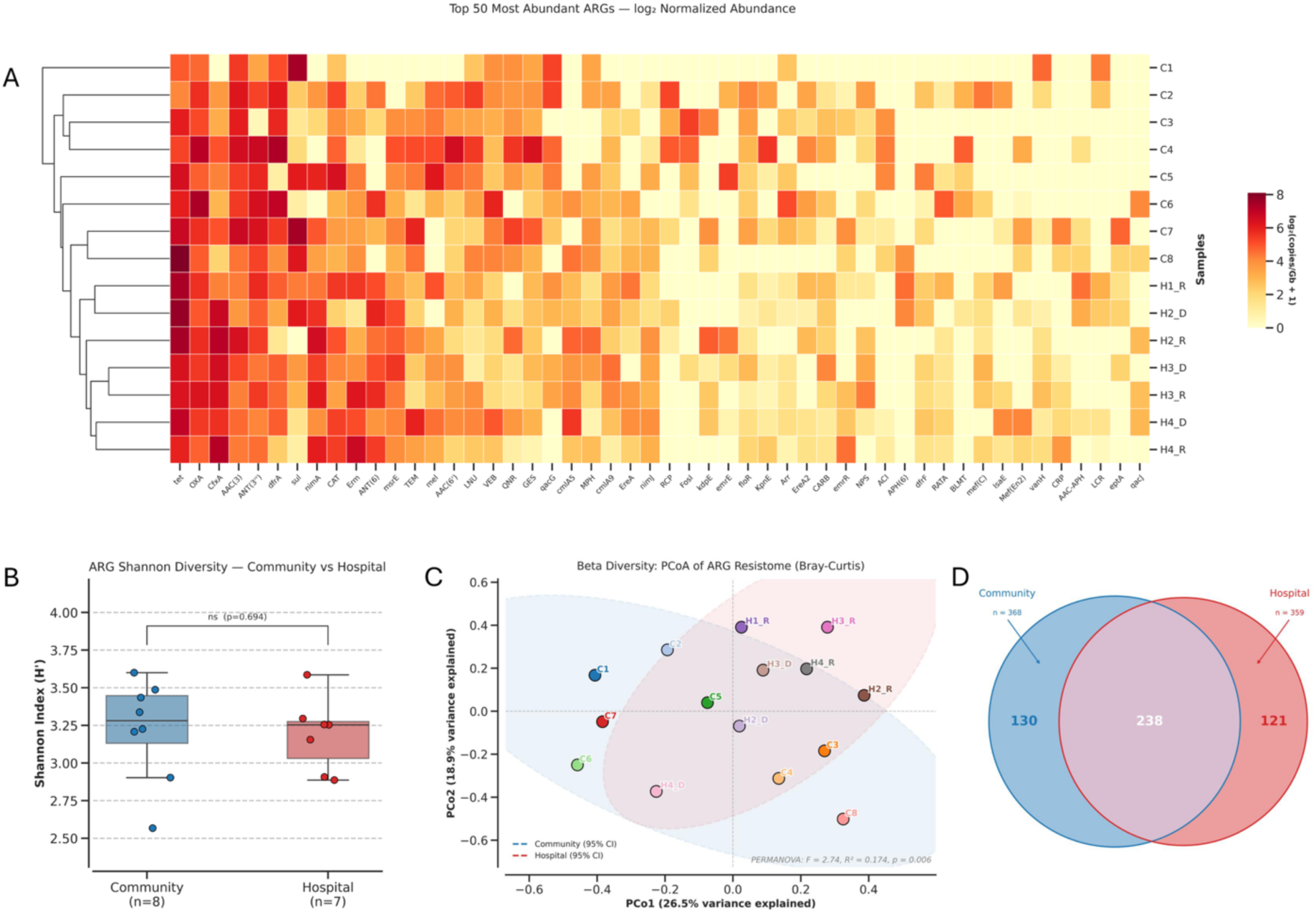
Resistome composition and dominant antimicrobial resistance genes identified in community and hospital wastewater samples from Conakry, Guinea. (A) Heatmap showing the normalized abundance (log₂-transformed copies/Gb) of the 50 most abundant ARGs detected across community and hospital wastewater samples.

Alpha diversity analysis revealed no significant differences in ARG diversity between community wastewater and hospital wastewater. The Shannon diversity index was comparable between community and hospital samples (p = 0.6943), indicating a similar level of ARG richness and evenness within both environments. Similarly, Simpson diversity values showed no significant variation between sources (p = 0.3969), suggesting an equally balanced structure of dominant ARGs. This absence of significant difference was consistent across sensitivity analyses restricting the hospital group to each individual sampling campaign (Shannon: p=0.461 for H_R, p=0.921 for H_D; Simpson: p=0.283 for H_R, p=0.921 for H_D). (**Figure 3B).** Contrary, principal coordinate analysis based on Bray-Curtis distance demonstrated a separation between community and hospital wastewater resistomes. PERMANOVA confirmed the sample source significantly influenced ARG composition (p = 0.006), explaining approximately 17.4% of the observed variance (**Figure 3C)**. We repeated the PERMANOVA separately against each hospital campaign. The separation remained significant against the rainy-season campaign (p=0.004) but was not statistically significant against the dry-season campaign (p=0.236).

Classification of ARGs according to antibiotic classes showed that genes associated with cephalosporins constituted the most abundant resistance group. Within this class, blaCfxA1 was the predominant determinant, followed by other members of the *bla*CfxA family (*cfxA2*, *cfxA3*, and *cfxA5*), as well as β-lactamases belonging to the *bla*TEM and *bla*OXA families. Genes associated with aminoglycoside resistance represented the second most abundant group and were mainly composed of determinants from the *aad, aac, aph*, and ant families, indicating a high diversity of aminoglycoside-modifying enzymes.

Among penicillin-associated β-lactam resistance genes, *bla*TEM-1 and *bla*OXA-family β-lactamases were the dominant determinants, followed by additional β-lactamase variants including members of the *blaGES* family. Tetracycline resistance genes were also highly represented, with *tetW* being the most abundant determinant, followed by *tetM, tetA, tetQ*, and several additional members of the *tet* family. Although less abundant than the major resistance classes, genes associated with resistance to critically important antibiotics were also detected. Carbapenem-associated ARGs were dominated by *bla*GES, followed by additional carbapenemase-related determinants detected at lower abundance. Furthermore, multidrug resistance genes, including efflux pump-associated determinants, were consistently identified across samples (**Figure 4)**.

**Figure 4:**
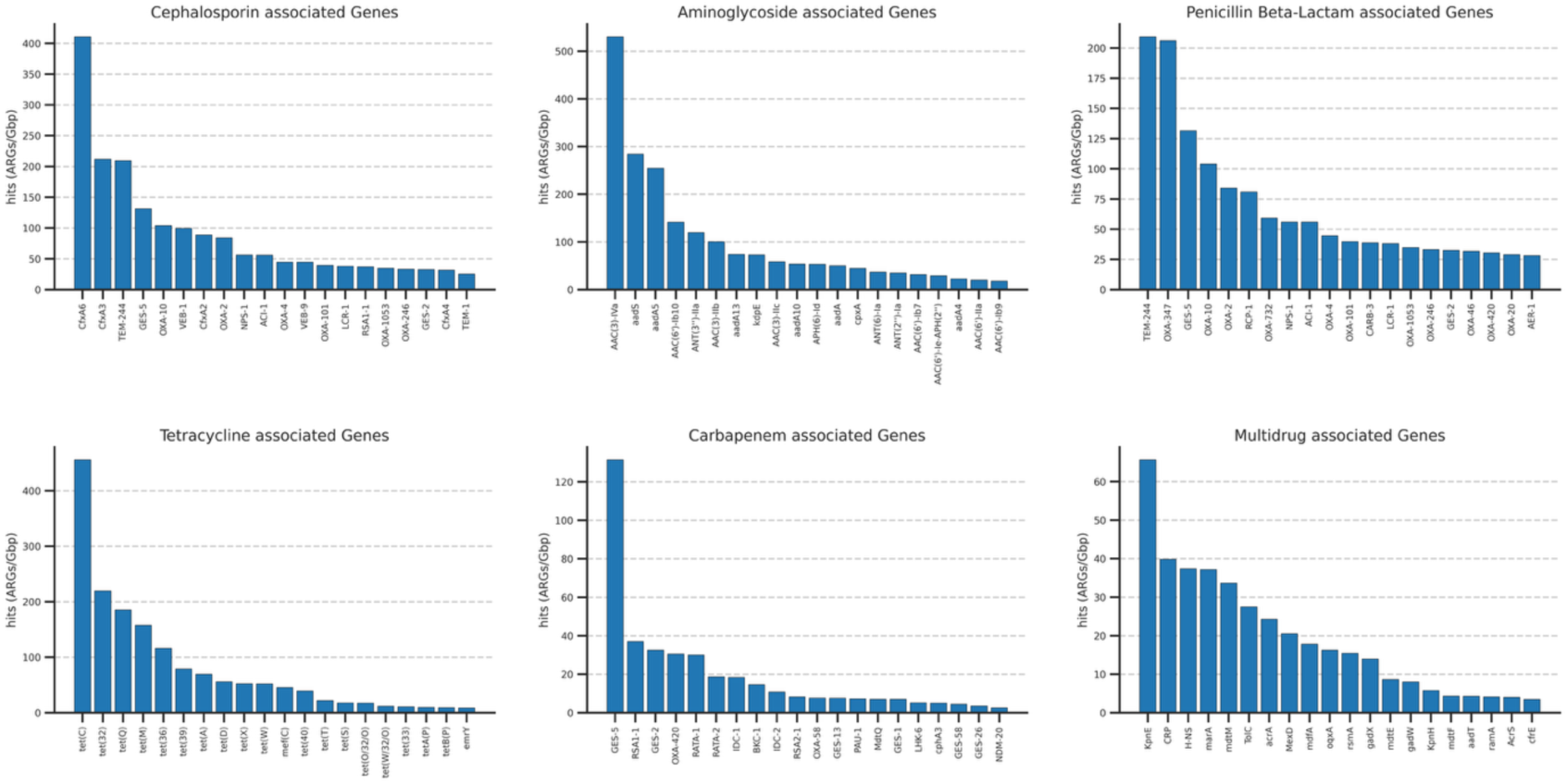
Most abundant ARGs associated with major antibiotic classes.

### 3.3. Antibiotic classes and Resistance mechanism

The overall relative distribution of ARG classes was similar across all wastewater samples, with resistance genes associated with β-lactams, aminoglycosides, tetracyclines, nitroimidazoles, and multidrug resistance representing the most abundant categories **(Figure 5A)**. While some variability was observed among individual samples, no major shifts in ARG class composition were evident between community and hospital wastewater. Resistance mechanisms were dominated by antibiotic inactivation, which accounted for approximately half of all detected ARGs across samples. Mechanisms related to antibiotic efflux, target protection, and target alteration, were also detected in all samples. **(Figure 5B)**

**Figure 5:**
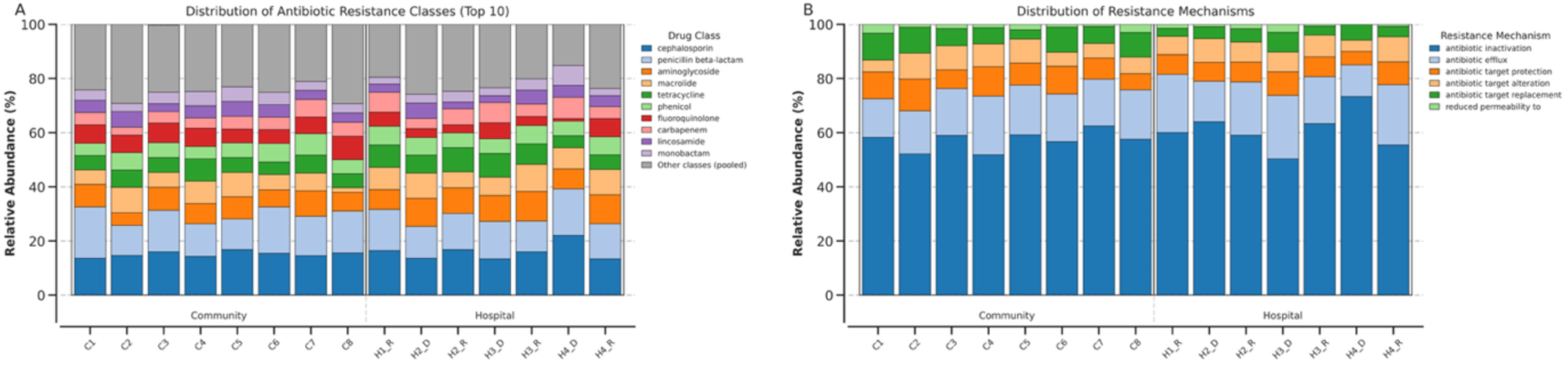
Distribution of antimicrobial resistance classes and resistance mechanisms across community and hospital wastewater samples from Conakry, Guinea. (A) Relative abundance of antimicrobial resistance genes (ARGs) grouped by antibiotic class (B) Relative distribution of resistance mechanisms

### 3.4. Mobile genetic elements

Mobile genetic element (MGE) profiling based on the mobileOG-db database identified the five major functional categories across all 15 wastewater samples **(Figure 6)**. Integration/excision elements were the predominant category, accounting for an average of 49.5% of all detected MGEs, followed by transfer elements (20.5%), replication/recombination/repair elements (11.8%), phages (9.1%), and stability/transfer/defense elements (9.2%). All five categories were detected in every sample (100% prevalence), highlighting the widespread occurrence of genetic mobility mechanisms within both community and hospital wastewater environments.

**Figure 6:**
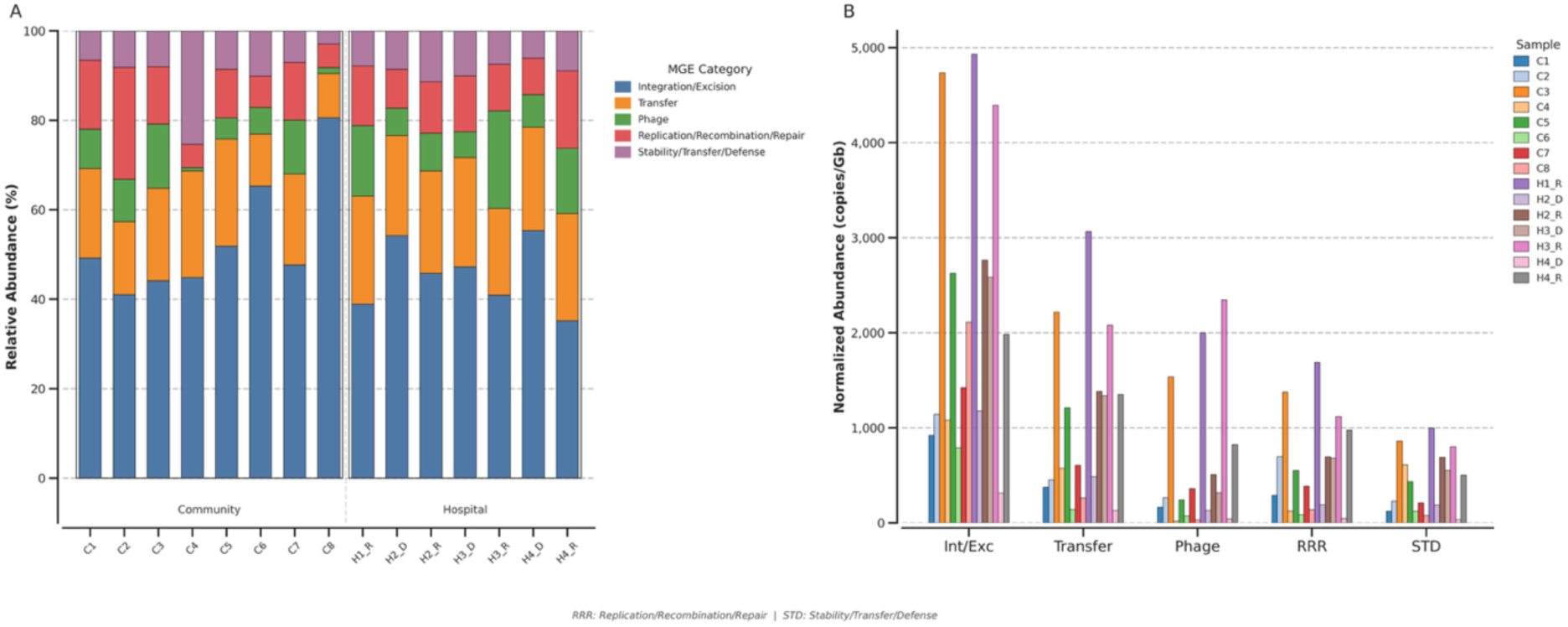
Composition and abundance of mobile genetic element (MGE) categories in community and hospital wastewater samples from Conakry, Guinea. (A) Relative abundance of the five major MGE functional categories identified using mobileOG-db, including integration/excision, transfer, phage, replication/recombination/repair (RRR), and stability/transfer/defense (STD), across community and hospital wastewater samples. (B) Normalized abundance (copies/Gb) of MGE categories in individual samples, highlighting variations in mobilome burden between community and hospital wastewater.

The relative composition of MGEs was broadly similar between community and hospital wastewater samples, although notable differences were observed. Community wastewater showed a numerically higher proportion of integration/excision elements (53.1% vs. 45.4%), whereas hospital wastewater showed higher proportion of transfer elements (22.9% vs. 18.3%) and phage-associated sequences (11.4% vs. 7.2%) **(Figure 6A)**. In contrast, categories associated with replication/recombination/repair (11.8%) and stability/transfer/defense (9%) showed relatively consistent proportions across both environments.

After normalization, the total MGE burden reached 72,979.7 copies/Gb, with a higher cumulative abundance in hospital wastewater (43,308.3 copies/Gb) than in community wastewater (29,671.3 copies/Gb). Integration/excision elements represented the largest fraction of the mobilome (32,979.6 copies/Gb), followed by transfer elements (15,672.3 copies/Gb), replication/recombination/repair elements (9,037.8 copies/Gb), phages (8,853.7 copies/Gb), and stability/transfer/defense elements (6,436.2 copies/Gb) **(Figure 6B)**.

PlasmidFinder analysis identified 54 plasmid replicon sequences representing 18 distinct incompatibility groups across the 15 wastewater assemblies. Replicons were detected in both hospital and community wastewater, with IncP6 being the most frequently identified incompatibility group, followed by members of the IncF family (*IncFIA, IncFIB* and *IncFII*), IncC, IncR and repUS43. Additional replicon types, including *IncN, IncQ1, IncQ2, IncX3, IncY, IncU, IncA, IncB/O/K/Z* and *p0111*, were detected at lower frequencies. IncF-family replicons were identified exclusively in hospital wastewater, whereas *IncP6, IncC* and *IncR* were detected in both wastewater sources **(Supplementary table S3)**.

### 3.5 Genome-resolved characterization of ARG-carrying bacteria

Genome-resolved metagenomic analysis reconstructed 103 dereplicated metagenome-assembled genomes (MAGs), including 101 bacterial and two archaeal genomes. Based on completeness and contamination estimates, 80 MAGs carried at least one antimicrobial resistance gene. Among these, 26 MAGs exhibited contig-level ARG–MGE co-localizations (≤10 kb), representing putative mobile resistance regions.

Taxonomic assignment showed that these MAGs belonged to both clinically relevant and environmental bacterial taxa, including *Escherichia coli, Citrobacter amalonaticus, Parabacteroides distasonis, Bacteroides graminisolvens, Trichococcus flocculiformis, Faecalibacterium longum, Prevotella spp*., and *Thauera sp*.

The *Escherichia coli* MAG contained the highest number of ARG–MGE co-localizations (n = 52), followed by *Thauera sp*. (n = 18), *Citrobacter amalonaticus* (n = 13), *Prevotella sp*. (n = 9), and *Brachymonas sp*. (n = 9). Most co-localizations involved integration/excision-associated genes, and transfer, phage, replication/recombination/repair, and stability/transfer/defense-related elements were also observed **(Supplementary Table S4)**.

## 4. Discussion

To assess the burden of antimicrobial resistance in Guinea at a given point in time through wastewater-based surveillance, we analyzed 15 samples collected across communal medical centers and surrounding neighborhoods in Conakry. As a densely populated capital marked by high human activity, the city provides a critical setting for environmental AMR monitoring.

Our results reveal a highly diverse wastewater resistome, characterized by the detection of a large number of unique ARGs and a high overall abundance. Although hospital effluents are traditionally regarded as the primary reservoirs and environmental sources of antimicrobial resistance, our data show that community wastewater harbored a comparable ARG richness and an equivalent normalized abundance. In addition, nearly half of the detected ARGs were shared between the two environments, constituting a core resistome. Despite significant differences in bacterial alpha diversity, the substantial overlap in ARG composition and abundance indicates that antimicrobial resistance is present throughout the urban wastewater network rather than being confined to healthcare facilities. Similar observations have been reported in several metagenomic studies, where municipal wastewater exhibited resistome profiles comparable to those of hospital effluents [3,11,29–31]. This pattern may be explained by the cumulative contribution of the fecal microbiota of the general population, widespread antibiotic self-medication, the frequent dispensing of antibiotics without prescription in community pharmacies, and the high level of antibiotic prescribing in hospitals and private clinics without prior microbiological testing [32].

Although the overall ARG abundance and richness were comparable between the two wastewater sources, their resistome composition differed significantly. The hospital resistome exhibited a more homogeneous profile, likely reflecting stronger and more specific antibiotic selection pressures associated with the intensive use of broad-spectrum and last-resort antibiotics in healthcare settings [6,8,13,33]. In contrast, the greater dispersion of community wastewater samples may reflect the heterogeneity and plasticity of this reservoir, which is influenced by multiple and diffuse sources of contamination [11,34].

The predominance of β-lactam resistance genes, together with aminoglycoside, tetracycline, nitroimidazole, and multidrug resistance determinants observed in our study, is consistent with the global composition of wastewater resistome reported in studies conducted across Europe, Asia, and Africa [6,10,12,35–38]. The associated antibiotic classes are among the most frequently prescribed in both hospital and community settings, particularly in resource-limited countries where antimicrobial stewardship programs are not yet fully implemented [32,39]. In particular, the predominance of cephalosporin-related ARGs, largely driven by the *bla*CfxA family, may reflect the substantial contribution of anaerobic gut bacteria, particularly members of the genus Bacteroides, which are recognized reservoirs of these β-lactamases in the human gut microbiota [16,29,35]. Similarly, the widespread occurrence of *bla*TEM and *bla*OXA genes highlights the broad dissemination of β-lactam resistance determinants in urban wastewater.

Genes encoding aminoglycoside-modifying enzymes represented the second most abundant resistance group, could reflected the broad diversity of enzymatic mechanisms involved in aminoglycoside inactivation [13,15,34,40]. Tetracycline resistance genes, consistently detected in both wastewater sources has often been attributed to the long history of tetracycline use in human and veterinary medicine, making tetracycline resistance genes among the most widespread ARGs reported in environmental ecosystems [35,41].

Although detected at lower abundance, ARGs associated with critically important antibiotics deserve particular attention. The presence of carbapenemase-associated genes, including members of the *bla*GES family, together with multidrug resistance determinants and efflux pump-associated genes, suggests that clinically relevant resistance determinants are already circulating in the Guinean wastewater environment. Even at low abundance, these determinants represent a significant public health concern, as wastewater ecosystems provide favourable conditions for bacterial interactions and horizontal gene transfer, potentially facilitating their dissemination into previously unexposed environmental and clinical bacterial populations [12,29,34,40].

In our study, several bacterial groups widely recognized as major contributors to the global antimicrobial resistance crisis, including members of the Enterobacteriaceae family and ESKAPE pathogens, were identified. The detection of these bacterial groups in both community and hospital wastewater confirms that these environments serve as important reservoirs of microorganisms potentially involved in the dissemination of antimicrobial resistance. These bacteria are characterized by remarkable genomic plasticity, enabling them to acquire, maintain, and disseminate antimicrobial resistance determinants through horizontal gene transfer [14,16,33]. Genome-resolved analyses further showed that ARGs were associated with both clinically relevant and environmental bacterial taxa.

Several of the identified species are recognized as important hosts of plasmids, integrons, transposons, and other MGEs involved in the dissemination of clinically important ARGs [16,37]. In resource-limited countries, where sanitation infrastructure remains inadequate, these findings are of particular concern. Insufficiently treated wastewater can contribute to the dissemination of pathogenic bacteria and their resistance determinants into surface waters and soils, thereby promoting their circulation at the human–animal–environment interface [12,18].

While hospital wastewater is often considered an important hotspot for antimicrobial resistance, our results demonstrate that community wastewater also harbours a wide diversity of resistance genes, clinically relevant bacterial taxa, mobile genetic elements, and plasmid replicons. This convergence suggests that AMR circulates throughout interconnected urban wastewater systems rather than remaining confined to healthcare facilities. This may serve as a warning to health policymakers that action must be taken to strengthen the monitoring of these ecosystems, and also to put in place a policy for sanitation and the appropriate management of wastewater.

Our results highlight the potential of wastewater as a tool for integrated monitoring of AMR in Guinea. In a context where microbiological surveillance relies primarily on clinical isolates, environmental metagenomics offers a complementary approach that captures a much broader fraction of microbial diversity and resistance genes.

Our findings also suggest that community and hospital wastewater provide complementary information. Whilst hospital wastewater reflects the associated selection pressures, community wastewater serves as an indicator of the circulation of resistance at the community level. Incorporating these two sources into a national surveillance system could therefore provide a more comprehensive picture of the dynamics of AMR in Guinea.

Nevertheless, several limitations should be acknowledged. This study included a limited number of sampling sites and samples, relied on single grab samples rather than composite sampling, and was based on a single community sampling campaign, whereas hospital sites were sampled during two seasonal campaigns; this asymmetry precluded a fully balanced seasonal assessment across both source types. Also, longitudinal sampling of both source types should be implemented in future work.

Despite these limitations, this study establishes the first genomic baseline for wastewater-based AMR surveillance in Guinea, demonstrating the feasibility of generating comprehensive environmental data through long-read metagenomics and genome-resolved analyses in a resource-limited setting. Our findings support the integration of wastewater-based surveillance as a complementary component of national One Health AMR monitoring programmes in Guinea and provide a foundation for future longitudinal studies integrating human, animal, and environmental reservoirs.

## Data availability statement

The authors confirm that the data supporting the findings of this study are available within the article and its additional information. All raw sequencing data will be available via the NCBI Sequence Read Archive under the Bio Project ID PRJNA1511345. The analysis scripts are shared in the publicly available in GitHub repository at https://github.com/armelgnimadi/Resistome

## Author contributions

**TACG**: Writing – original draft, Conceptualization, Visualization, Software, Methodology, Investigation, Formal analysis, Data curation. **AkK**: Writing–review & editing, Methodology. **YMGH**: Writing–review & editing, Methodology, Validation **KEA**: Writing–review & editing, Methodology, Investigation, Data curation. **JFZ**: Supervision, Resources, Project administration. **AT**: Writing–review & editing, Supervision, Conceptualization, Project administration, Validation, Resources, Funding acquisition **MJM**: Writing – review & editing, Supervision, Methodology, Software, Validation, Resources. **AKK**: Writing – review & editing, Supervision, Resources, Project administration, Methodology, Investigation, Funding acquisition, Conceptualization.

## Acknowledgments

We would like to thank our partner TransVIHMI (IRD) for their logistical support, AFROSCREEN project, DOPERAUS project. And the Embassy of France in Guinea for funding the thesis through which this work was conducted.

