## Supplemental Table 4 for "Long-read metagenomics reveals a high burden of antimicrobial resistance, mobile genetic elements and bacterial diversity in hospital and community wastewater from Conakry, Guinea"

Table 1: All MAGs with confirmed ARG-MGE co-localization

Genome-resolved analysis, contig-level colocalization (<=10 kb), all taxa

| MAG ID | Taxonomic assignment | Completeness (%) | Contamination (%) | ARG(s) detected | MGE categor(y/ies) associated | N co-occurrence pairs |
| --- | --- | --- | --- | --- | --- | --- |
| MAG01 | *Escherichia coli* | 100.0 | 0.2 | acrB, AcrF, bacA, emrB, acrA, ampC β-lactamase, mdfA, H-NS, kdpE, marA, mdtA, mdtB, mdtC, mdtE, mdtF, mdtG, mdtH, mdtM, msbA, PmrF, rsmA, TolC, ugd, YojI | integration/excision, phage, replication/recombination/repair, stability/transfer/defense, transfer | 52 |
| MAG02 | *Thauera sp.* | 86.1 | 2.0 | APH(3'')-Ib, APH(6)-Id, sul1 | integration/excision, stability/transfer/defense | 18 |
| MAG03 | *Citrobacter amalonaticus* | 100.0 | 4.6 | bacA, acrA, H-NS, kdpE, mdtG, msbA, rsmA | integration/excision, phage, replication/recombination/repair, transfer | 13 |
| MAG04 | *Prevotella sp.* | 100.0 | 0.2 | CfxA3, Mef(En2) | integration/excision, transfer | 9 |
| MAG05 | *Brachymonas sp.* | 87.7 | 2.8 | mphE, msrE, tet(C) | integration/excision | 9 |
| MAG06 | *Desulfomicrobium sp.* | 57.0 | 0.9 | aadA, OXA-2 | integration/excision | 8 |
| MAG07 | *Macellibacteroides fermentans* | 92.9 | 2.6 | ErmF, ErmG, OXA-347 | integration/excision | 7 |
| MAG08 | *Desulfobulbus sp.5* | 99.2 | 7.2 | sul2 | integration/excision, stability/transfer/defense, transfer | 5 |
| MAG09 | Unclassified | 71.5 | 6.6 | lnuB, lsaE | integration/excision | 4 |
| MAG10 | *Bacteroides graminisolvens* | 83.0 | 8.4 | lnuB, lsaE, tet(Q) | integration/excision | 4 |
| MAG11 | *Trichococcus flocculiformis* | 100.0 | 4.3 | AAC(6')-Ie-APH(2'')-Ia bifunctional protein, ANT(6)-Ia | integration/excision | 4 |
| MAG12 | *Propionivibrio sp.* | 69.9 | 3.4 | qacEdelta1, sul1 | integration/excision, stability/transfer/defense | 4 |
| MAG13 | *Phocaeicola sp.* | 56.1 | 1.4 | CfxA3, sul2 | integration/excision, transfer | 4 |
| MAG14 | *Alloprevotella sp.* | 92.0 | 1.7 | CfxA3, nimA | integration/excision, transfer | 4 |
| MAG15 | *Lachnospira rogosae* | 71.4 | 2.2 | tet(O) | integration/excision, transfer | 4 |
| MAG16 | *CAYUJY01 sp.* | 86.1 | 0.1 | Erm(A), catQ | integration/excision | 3 |
| MAG17 | *Parabacteroides distasonis* | 70.3 | 0.7 | CfxA3, tet(Q) | stability/transfer/defense, transfer | 3 |
| MAG18 | *AWVT01 sp.* | 83.1 | 2.1 | tet(32) | replication/recombination/repair, transfer | 2 |
| MAG19 | *UBA7485 sp.* | 98.8 | 1.6 | tet(O) | transfer | 1 |
| MAG20 | *Sulfurospirillum sp.* | 97.0 | 0.1 | lnuC | integration/excision | 1 |
| MAG21 | *MWDD01 sp.* | 79.4 | 2.2 | EreA | integration/excision | 1 |
| MAG22 | *Saccharofermentans sp.* | 96.4 | 2.7 | ANT(6)-Ia | integration/excision | 1 |
| MAG23 | *Alloprevotella timonensis* | 83.4 | 5.4 | CfxA3 | transfer | 1 |
| MAG24 | *Faecalibacterium longum* | 84.8 | 2.9 | tet(W) | integration/excision | 1 |
| MAG25 | *Alloprevotella sp.* | 87.3 | 2.9 | tet(Q) | integration/excision | 1 |
| MAG26 | *Alicycliphilus limosus* | 59.3 | 5.9 | aadA5 | integration/excision | 1 |
