## Supplemental Table 1 for "Long-read metagenomics reveals a high burden of antimicrobial resistance, mobile genetic elements and bacterial diversity in hospital and community wastewater from Conakry, Guinea"

Table 1: Raw sequencing statistics per sample

| Sample | N reads | Total bases (Gb) | Mean read length (bp) | N50 read length (bp) | Mean quality (Q) |
| --- | --- | --- | --- | --- | --- |
| Community*^1^* | | | | | |
| C1 | 2,747,491 | 2.58 | 938 | 2,260 | 15.2 |
| C2 | 1,008,835 | 1.08 | 1,069 | 2,463 | 15.5 |
| C3 | 664,940 | 1.80 | 2,708 | 5,429 | 15.3 |
| C4 | 113,268 | 0.29 | 2,538 | 5,496 | 15.2 |
| C5 | 774,142 | 1.71 | 2,213 | 5,629 | 15.3 |
| C6 | 2,073,058 | 1.40 | 676 | 1,177 | 15.4 |
| C7 | 1,876,902 | 1.81 | 965 | 2,682 | 15.7 |
| C8 | 88,134 | 0.21 | 2,360 | 4,459 | 15.1 |
| Hospital | | | | | |
| H1_R | 1,552,394 | 1.97 | 1,270 | 4,641 | 16.7 |
| H2_D | 471,696 | 1.46 | 3,104 | 5,691 | 15.3 |
| H2_R | 397,829 | 1.38 | 3,457 | 7,660 | 16.7 |
| H3_D | 638,720 | 1.15 | 1,805 | 4,732 | 15.5 |
| H3_R | 981,073 | 3.47 | 3,535 | 7,211 | 16.3 |
| H4_D | 2,251,883 | 3.16 | 1,404 | 3,322 | 15.1 |
| H4_R | 1,117,484 | 2.37 | 2,120 | 5,484 | 16.3 |
| *^1^*C4 and C8 were excluded from genome-resolved (MAG) analyses due to insufficient sequencing depth. | | | | | |
